# Expanding the mutational spectrum of *GCK* in Turkish pediatric population

**DOI:** 10.1101/2024.02.07.24302418

**Authors:** Vehap Topcu, Gonul Buyukyilmaz, Keziban Toksoy Adiguzel, Mehmet Boyraz

**Affiliations:** Department of Medical Genetics, Ankara City Hospital, Ankara, Turkey; Department of Pediatric Endocrinology, Ankara City Hospital, Ankara, Turkey; Department of Pediatric Endocrinology, Ankara Yildirim Beyazit University Faculty of Medicine Ankara, Turkey

**Keywords:** glucokinase, hyperglycemia, GCK-MODY, monogenic diabetes, pediatric patients

## Abstract

Heterozygous loss-of-function variants in *GCK* cause persistent, mildly elevated plasma glucose beginning at birth. Recently, clinical phenotype created by deleterious variants in *GCK* is called GCK-MODY, although currently registered as “MODY, type 2” (maturity-onset diabetes of the young type 2, MODY2, MIM # 125851) in OMIM. The hyperglycemia of GCK-MODY is a benign and non-progressive condition, usually suspected upon detection of hyperglycemia in adulthood. Medication is not essential, nevertheless, pregnant females may require special attention. Here, we report recurrent and novel *GCK* variants detected in 43 pediatric patients who were investigated for hyperglycemia by the referring pediatric endocrinologist. Electronic medical records (EMR) of the patients were collected and reviewed retrospectively. All patients applied to Ankara City Hospital between April 2019 and June 2022. *GCK* variants were screened on NovaSeq 6000 next-generation sequencing platform (Illumina). Variants detected in *GCK* were checked with recent literature for a proper pathogenicity classification. In 43 patients, 28 distinct *GCK* variants were identified. Of these variants, 25 were classified as likely pathogenic/pathogenic variants (c.1342G>A, c.112C>T, c.1178T>C, c.130G>A, c.565A>G, c.208+3A>T, c.349G>C, c.863+5G>A, c.214G>A, c.1292C>A, c.830_831delTG, c.106C>T, c.572G>A, c.107G>C, c.454T>C, c.793G>A, c.645C>A, c.377T>A, c.667G>A, c.46-1G>A, c.149A>C, c.1079C>G, c.401T>C, c.758T>G, c.467_483+6del; NM_000162.5), while remaining 3 variants (c.950A>C, c.186G>T, c.188G>A; NM_000162.5) were classified as variant of uncertain significance. According to the variant effect, missense variants accounted for the majority (71%; 20/28), followed by splice junction (14%; 4/28), premature termination (7%; 2/28), frameshift (4%; 1/28), and synonymous (4%; 1/28) alterations. There were 3 novel variants: c.830_831del, c.377T>A, c.467_483+6del.

## INTRODUCTION

First evidence of monogenic diabetes emerged from the study that suggested a role of glucokinase locus in French MODY families (Froguel et al., 1992; Froguel et al., 1993). Clinical phenotype caused by heterozygous inactivating variants in glucokinase (GCK) enzyme is recently named GCK-MODY, also known as MODY2 (OMIM entry #125851). Frequency of pathogenic variants in glucokinase varies in different geographical regions and comprises of 30% to 60% of all MODY phenotypes (Shields et al., 2010). Turkish MODY series figured a similar frequency, ranging between %24-%60 (Aydogan et al., 2022). Albeit a prevalence of 1 in 1000 individuals is proposed (Chakera et al., 2015), a certain proportion of the GCK-MODY is hidden likely because of misdiagnosis or coincidence with type 1 or type 2 diabetes (Martin et al., 2008). It’s prevalence among all diabetes patients is around 2-5% worldwide.

*GCK* is copiously expressed in the pancreas, hepatocytes, and brain tissues. It tunes insulin secretion in response to alterations in blood glucose levels, thus keeping serum glucose levels over a physiological range. The cellular expression of GCK isoforms is controlled by different enzymes, regulators, and cellular substances. While the islet-specific isoform is regulated directly by glucose, the liver-specific isoform is under the control of both insulin and glucokinase regulatory protein (GKRP) (Choi, Seo, Kyeong, Kim, & Kim, 2013; Matschinsky, 1990). Monoallelic loss of function variants is sufficient to exhibit disease features. Over 1000 hundred mutations are registered in the Human Gene Mutation Database (https://portal.biobase-international.com/hgmd/pro/start.php), scattering evenly along the exons and exon-intron boundaries.

A mild fasting hyperglycemia presenting right after birth is the hallmark of GCK-MODY (Prisco, Iafusco, Franzese, Sulli, & Barbetti, 2000); however, in older ages it is underdiagnosed because of the lack of classical symptoms and diabetic complications (Steele et al., 2014). Frequently, GCK-MODY is uncovered in a routine medical investigation. A fasting plasma glucose (FPG) between 5.4 and 8.3 mmol/L and glycated hemoglobin A1c (HbA1c) between 5.8 and 7.6% (40–60 mmol/mol) are in favor of GCK-MODY diagnosis (Chakera et al., 2015). Oral glucose tolerance test reveals a trivial incremental elevation of the blood glucose and a mild increase in A1c (HbA1c) (Steele et al., 2013). Accurate diagnosis is crucial to differentiate GCK-MODY from early type 1 and type 2 diabetes for a convenient counseling and to pick appropriate course of management and surveillance of the patients and family members with GCK-MODY. Today, clinical diagnosis is confirmed conveniently using widely available molecular genetic testing, mostly in adults (Chakera et al., 2015; Vionnet et al., 1992). Molecular genetic testing in GCK-MODY yields a high diagnostic rate, particularly in pedigrees with a dominant inheritance. Persistent fasting hyperglycemia remains stable throughout life, irrespective of mutation type. So, treatment is scarcely recommended as microvascular and macrovascular complications are not expected in GCK-MODY; however, pregnancy is an exception for which insulin treatment is indicated if the fetus has macrosomia in second-trimester fetal ultrasound (American Diabetes, 2020). Thus, detection of macrosomia may be an indicator of a fetus without an inherited pathogenic GCK variant.

Here, we report 28 distinct *GCK* variants observed in a series of 43 patients in Ankara City Hospital, Medical Genetics Laboratory referred by Clinic of Pediatric Endocrinology. We have identified published and novel variants in *GCK*, some of which have not been reported in Turkish GCK-MODY patients.

## MATERIALS AND METHODS

### Patients

In this retrospective study, we screened our in-house database to find individuals with a reported *GCK* variant. We included only patients referred from pediatric endocrinology outpatient clinic of Ankara City Hospital (Ankara, Türkiye) between 2019 April – 2022 June, totally 43 patients (27 female, 16 male) aged 0-18 years old. All patients had a clinical profile consistent with GCK-MODY. The following clinical and laboratory variables were collected using EMR: gender, age, reason for consultation, body mass index (BMI), and laboratory investigations including glycosylated hemoglobin A1c, FPG, 2-hour plasma glucose (2h-PG), fasting insulin (Fins) concentration, and 2-hour insulin concentration (2h-Ins). All patients were negative for anti-glutamic acid decarboxylase, anti-islet cell, and anti-insulin antibodies, thus type 1 diabetes were ruled out.

### Genetic analysis and variant classification

Genomic DNA was isolated from peripheral blood using QIAamp DNA mini kit (Qiagen, Hilden, Germany). We screened all exons and ± 20 bp exon-intron boundaries of *GCK* using Next Generation Sequencing (NGS). Sequencing libraries were prepared with CDHS-14622Z-506 QIAseq Targeted DNA Custom Panel (Qiagen, Hilden, Germany) and sequenced on Illumina NovaSeq 6000 platform (Illumina, San Diego, CA, USA). Read mapping and variant calling were performed using CLC Genomics Workbench (CLC-GWB, Qiagen). Variant analysis was performed on QIAGEN Clinical Insight (QCI) interpret software (QIAGEN, Hilden, Germany). Integrative Genome Viewer (IGV) was used for the visual analysis of depth coverage, sequences quality and variants identification. Next generation sequencing (NGS) covered 100% of the bases with more than 30x coverage.

*GCK* variants were reported with reference to the canonical transcript NM_000162.5 (β-cell isoform). Variants were classified according to the ACMG/AMP international guidelines for interpreting sequence variants (Richards et al., 2015), a system that helps assign a variant one of five classification categories: Pathogenic (P) ;likely pathogenic (LP), variant of uncertain significance (VUS), likely benign (LB), or benign (B). In the present study, we solely reported variants classified as pathogenic/likely pathogenic or uncertain significance. We searched all variants in dbSNP, ClinVar, HGMD, and medical literature; a *GCK* variant was deemed de novo if it was not registered in one of these variants databases or in published literature.

### Statistical analysis

All statistical analyses were carried out with the Statistical Package for Social Sciences (SPSS 15.0 for Windows, Chicago, IL, USA).

### Ethics approval

A written informed consent of patients and/or his/her parents is obtained prior to routine genetic testing. Retrospective analysis of the patient data was approved by the ethics committee of Ankara City Hospital (E2-22-2102). This research was conducted in accordance with 2013 Declaration of Helsinki.

## RESULTS

### Clinical and Biochemical Characteristics

Clinical data of the patients are summarized in (**Table 1, Supplementary** Figure 1). GCK-MODY does not show considerable clinical variability. The biochemical characteristics of our patients were comparable to those reported previously.

**Table 1.** General characteristics of the patients.

|  | No. of patients | Mean | ± SD |
| --- | --- | --- | --- |
| FPG (mg/dL) | 43 | 116,3 | 15,9 |
| 2h-PG (mg/dL) | 32 | 148,1 | 37,5 |
| HbA1c (%) | 43 | 6,4 | 0,5 |
| Fins (mU/L) | 42 | 7,9 | 7,7 |
| 2h-Ins (mU/L) | 30 | 35,1 | 39,8 |
| C-peptide (µg/L) | 41 | 1,0 | 0,6 |

Data is expressed as mean ± SD. FPG: Fasting plasma glucose; 2h-PG: 2-hour plasma glucose; Fins: Fasting insulin; 2h-Ins: 2-hour insulin.

The mean age of the patients was 9.3 years old. The gender distribution was 27 females (63%) and 16 males (37%) (**Figure 1**). The main reason for the consultation with medical genetics to get *GCK* sequenced was the laboratory investigation of persistent fasting hyperglycemia in all but two patients, who had polydipsia, polyuria, and polyphagia as primary complaints. Anthropometric measurement was available in 32 patients; the proportion of the patients for those with normal weight (84,4%), below-normal weight (6,3%), obesity (6,3%), and overweight (3,1%) was determined according to BMI.

**Figure 1.**
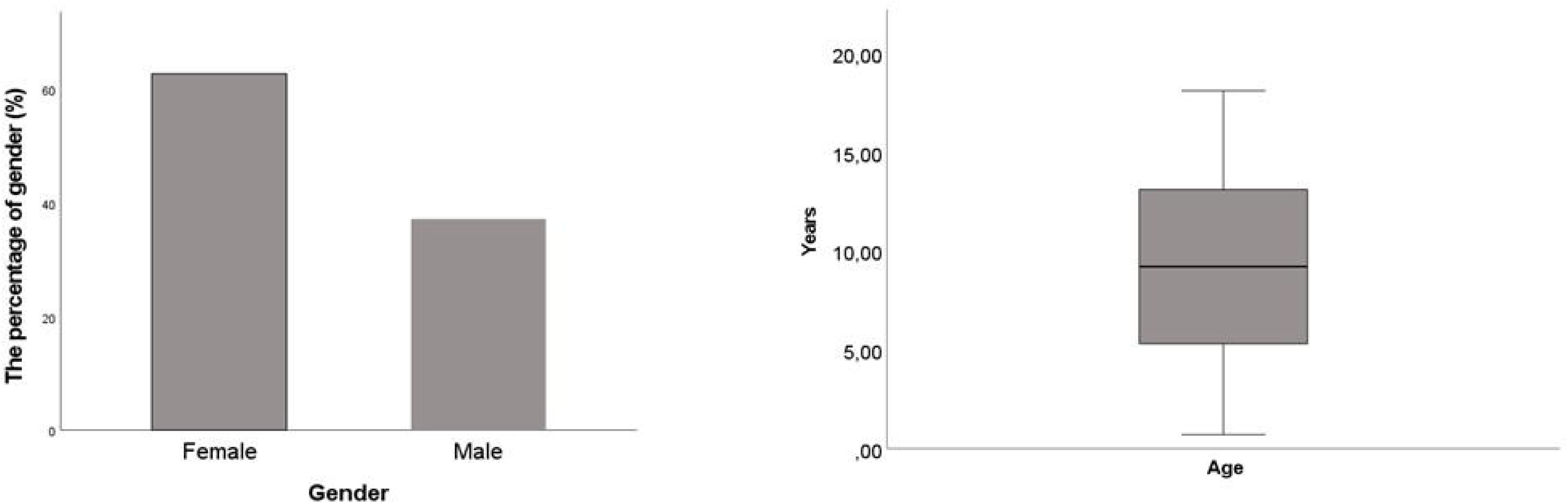
Gender and age distribution of the patients. A) Bar graph for gender distributiion and B) Whisker plot for age distribution.

All patients had a glycosylated hemoglobin A1c<%8 and were found negative for type 1 diabetes-related autoantibodies against glutamic acid decarboxylase (GAD), islet cells, and insulin. Medical history revealed coeliac disease in one patient (GCK-11), and Hashimoto’s disease (GCK-22) in another. None of the patients had hypertension, acanthosis nigricans (data was available in 33 of 43), microvascular complications, or diabetic ketoacidosis (DKA). There was not any patient on medication.

### *GCK* Variants

We observed 28 distinct variants in 43 patients, of these, 17 were classified as pathogenic (P), 8 were classified as likely pathogenic (LP), and 3 were classified as uncertain significance (VUS) according to the ACMG classification system (Richards et al., 2015). According to the effect on the transcript, there were 20 missense (71,4%) variants, followed by 4 splice site/near-splice-site (14,3%), 2 premature termination (7,1%), 1 frameshift (3,6%), and 1 synonymous (3,6%) alterations (**Table 2**). So, the majority of the GCK variants were detected in the coding sequence and uniformly distributed along the protein (**Figure 2**). We checked all variants from dbSNP, ClinVar, and HGMD databases, as well as searched medical literature (PubMed and Google Scholar). We regarded a variant as novel if it was not recorded in any of the databases or medical literature. We have observed 3 novel variants (c.830_831delTG, c.377T>A, c.467_483+6del), while the remaining 25 variants in this series were reported elsewhere. Four variants (c.1178T>C, c.208+3A>T, c.112C>T, c.1079C>G) were observed in more than one patient in this series (**Supplementary Table 1**).

**Figure 2.**
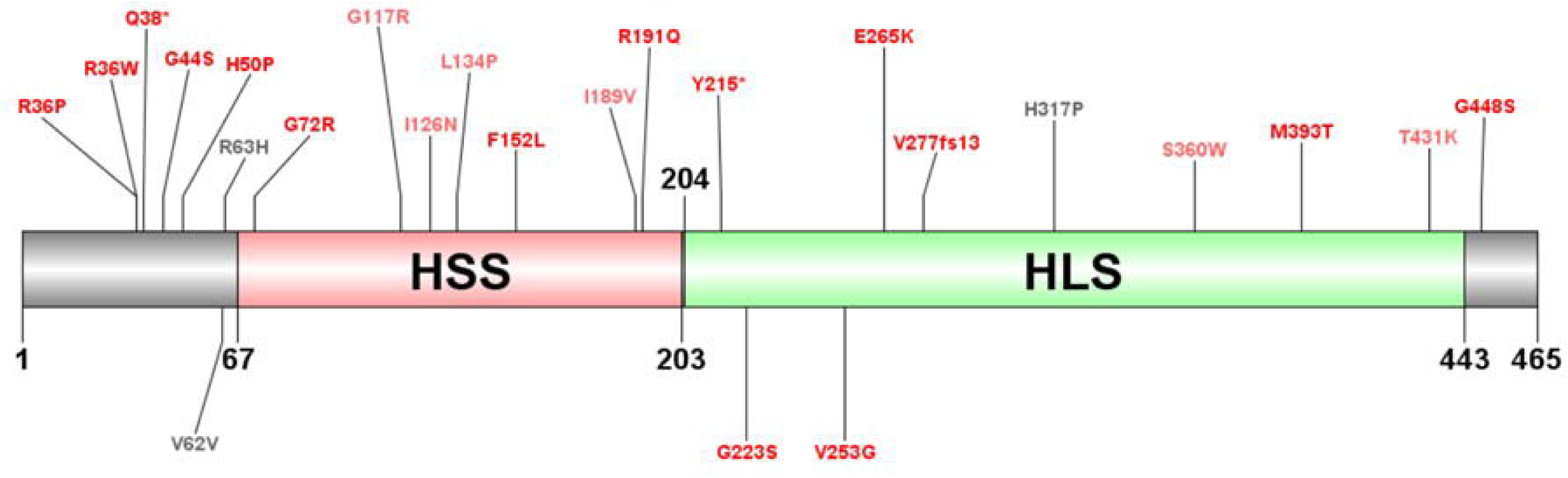
Distribution of the variants on Glucokinase. HSS, hexokinase small subdomain; HLS, hexokinase large subdomain. Variants are color­ coded: pathogenic, red; likely pathogenic, light red; variant of uncertain significance, gray.

**Table 2.**
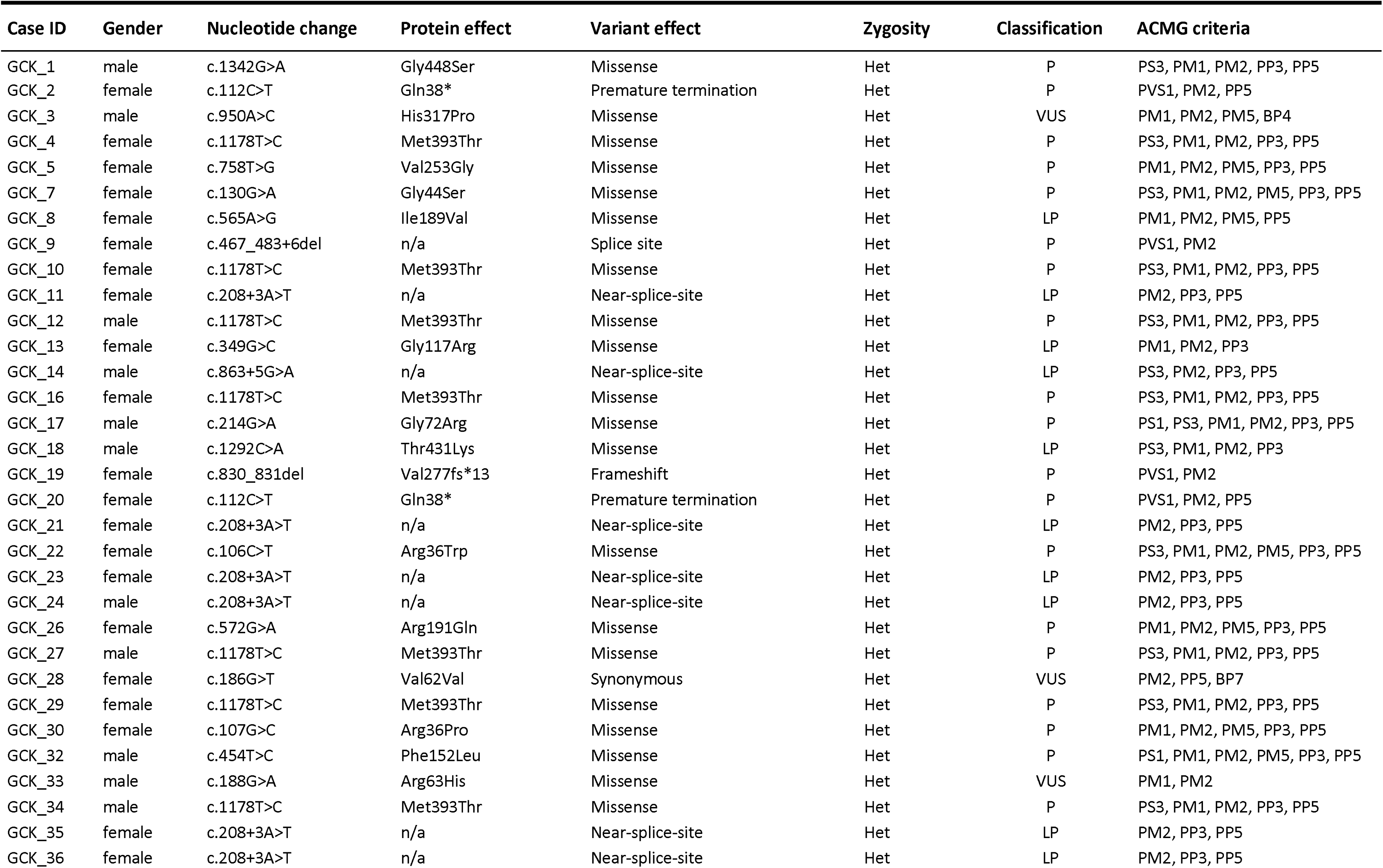
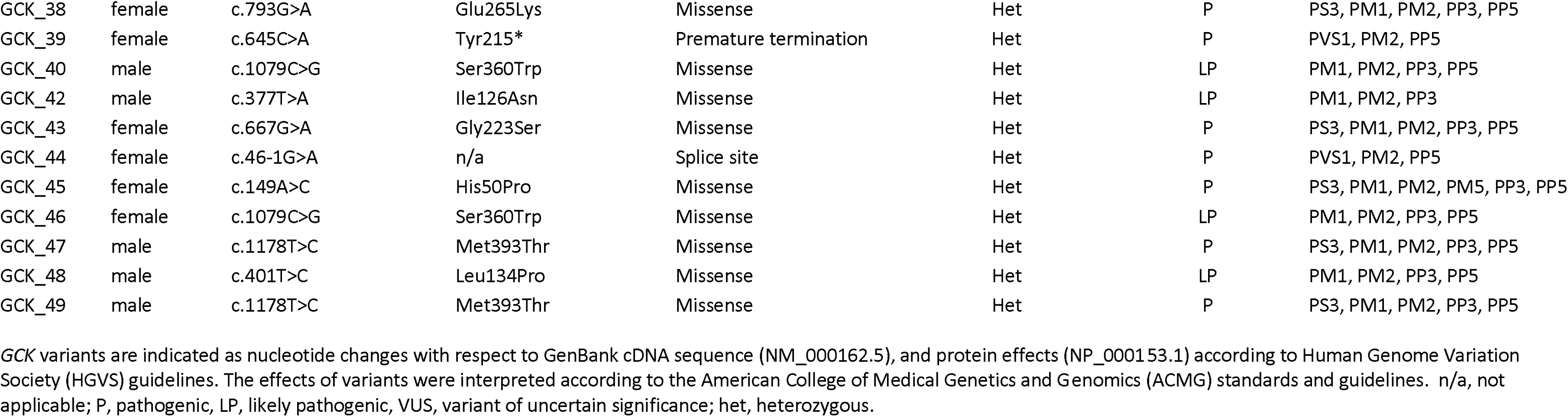
GCK variants detected in patients.

## DISCUSSION

We present the mutational spectrum of 43 pediatric patients referred for *GCK* sequencing to confirm the clinical diagnosis of GCK-MODY, the most prevalent form of monogenic diabetes. Thus, genetic analysis revealed 28 distinct rare *GCK* variants in our patient series, each patient having a single *GCK* alteration in one allele. Main complaint of the patients was fasting hyperglycemia and mild increase in the 2h-PG level revealed in routine or incidental blood glucose testing, while lacking classic signs of type 2 diabetes. Although GCK-MODY has an insidious onset, it has a high diagnosis rate before age 25; in agreement, ∼55% of our patients were diagnosed molecularly in the first decade. Awareness of this entity among pediatric endocrinologists and accessibility to DNA tests fostered prompt molecular diagnosis in younger ages.

Several Turkish patient series of GCK-MODY have been published previously, which figure out an approximate landscape of the GCK variants in the Turkish population (Agladioglu et al., 2016; Altan et al., 2020; Anik et al., 2015; Ates et al., 2021; Aykut et al., 2018; Bolu, Eroz, Dogan, Arslanoglu, & Dundar, 2020; Dundar, 2023; Duzkale & Emiroglu, 2023; Haliloglu et al., 2016; İşleyen & Semih, 2019; Ozdemir et al., 2018; Sağsak, Önder, Kendirci, Yıldız, & Doğan, 2022; Siklar & Berberoglu, 2016; Yalcintepe et al., 2021; Yilmaz Uzman et al., 2022). Still, only 58% of the variants detected in our patients overlapped with those in the previous series of the Turkish population. Therefore, 42% (12/28) of the variants we observed in our patients were reported for the first time in Turkish patients (**Supplementary Table 1**). In this regard, our report would further reflect the geographical view of *GCK* variants.

GCK encodes for 465 amino acid-length Hexokinase that consists of two subdomains, namely, hexokinase small subdomain (HSS, 67-203) and hexokinase large subdomain (HLS, 204-443). Over 1000 variants are registered in the Human Gene Mutation Database (Adzhubei et al., 2010), and there is no hot spot region or group of mutations to screen as a first-tier step for GCK-MODY *(Osbak et al., 2009).* In conformity, mutations detected in our patients tended to distribute along the exons and exon-intron junctions of the GCK uniformly; nevertheless, they were slightly more populated relatively in HSS compared to HLS concerning the domain size (Figure. Distribution of the variants on GCK). It was noteworthy that variants c.1178T>C (9 patients), c.208+3A>T (6 patients), c.112C>T (2 patients), c.1079C>G (2 patients) were observed in more than two cases, which, we think, is a chance occurred in our patient group, or these mutations may be more common in general. Regarding variant effect, most (71.4%) were missense type, while the remaining (28.6%) were causing premature termination, frameshift, synonymous events, or splice region alterations with possible harmful effects on proper splicing. Of these variants, 3 were novel.

Variant classification guidelines mention scrutinizing for experimental data of any variant in literature, which helps a more thorough and confident classification. The functional consequence of sequence variants on glucokinase activity is evaluated through protein stability and enzyme kinetic assays. Our comprehensive literature search yielded research papers including functional evidence for 11 of 28 variants detected in our patients, which prompted us to assign the PS3 criterion of the ACMG system (**Supplementary Table 1**). Functional evaluation of c.863+5G>A splice variant (GCK_14) was carried out using a minigene assay, which showed the deletion of exon 7 (Bouvet et al., 2023). In addition, regarding H50P alteration (GCK_45), there was no first-hand functional data; however, a nonsynonymous alteration at the same position (H50D) was experimented with by Raimondo et al. (2014), which demonstrated altered protein stability and kinetics. Thus, we deduced that H to P alteration would be damaging to the enzyme function given that Proline (P) does not have an electrically charged side chain and is smaller in size compared to Histidine (H), namely two amino acids have different molecular properties. Likewise, we did not find any direct functional evidence for R191Q but saw R191W was experimented with by Wang et al. (2019). Consequently, both H50D and R191Q were given PM5 instead of PS3, nevertheless, either variant ensured adequate points to be classified as pathogenic. Other variants with a functional study were Arg36Trp (Osbak et al., 2009), Gly44Ser (Wang et al., 2019), Gly72Arg (Raimondo et al., 2014), Gly223Ser (Valentinova et al., 2012), Glu265Lys (Osbak et al., 2009), Met393Thr (Raimondo et al., 2014), Thr431Lys (Lin et al., 2019), and Gly448Ser (Li et al., 2022).

In GCK_26, we observed a rare synonymous variant (c.189G>T, Val63Val), annotated as a disease-causing variant according to the Human Gene Mutation Database (HGMD). c.189G>T is a rare variant (ƒ = 0.00000398 in the gnomAD Exomes); however, splice predicting algorithms predicted no impact on the splice consensus sequence nor the creation of a new splice site (**Supplementary** Figure 2). Thus, we preferred to classify it as a likely benign/uncertain significance based on PM2 and BP4/BP7 criteria of the ACMG classification (Richards et al., 2015). To the best of our knowledge, there is no functional study assessing the splice effect of this variant.

## Conclusion

As Sanger sequencing and NGS became widespread and readily available for most healthcare facilities at a low cost, more healthcare practitioners now pick direct genetic testing based on suspicion of GCK-MODY, refraining from unnecessary laboratory investigation. The lack of hotspot variant(s)/region in *GCK* makes it credible to sequence all coding exons. One may prefer to analyze merely *GCK* or in a panel made up of prevalent monogenic diabetes genes (HNF4A, HNF1A, HNF1B, etc.). Given that GCK-MODY is straightforwardly distinguished from other monogenic forms, the NGS gene panel approach is not a requisite (Carroll & Murphy, 2013). In GCK-MODY, the patient and his/her family are informed that GKC heterozygotes do not require medication nor a surveillance program, whereas homozygous individuals present a severe outcome, known as Permanent neonatal diabetes mellitus-1 (MIM # 606176), which demands insulin treatment soon after birth. We consider it wise to communicate this piece of information to the parents and relatives, particularly when families are from geographical regions where endogamy is common.

Our study has some limitations. Retrospective data is obtained from a small group of pediatric patients with pathogenic GCK variants, so is not a broad representation of all possible *GKC* variants in the Turkish population.

The lack of segregation study of a *GCK* variant in a family was unfavorable in terms of exploiting ACMG criteria such as PP1 (co-segregation in affected family members) and PS2 (de novo variant). Nevertheless, the majority of the variants observed in this study fulfilled sufficient evidence to be LP/P even in the absence of PP1 and PS2. Variants H317P, V62V, and R63H remained uncertain significance as of now since we could not employ corresponding ACMG criteria relating to family study.

In most of our patients, information regarding the family history of individuals with diabetes was missing, so we did not have respectable family history data to provide. We are aware that a family history of diabetes at a young age (usually before 30 years of age) and a pedigree suggestive of monogenic diabetes are useful criteria along with biochemical evaluations in selecting patients before genetic testing. In our study, pediatric endocrinologists opted for *GCK* sequencing mainly resting on biochemical evidence.

## Author contribution statement

V.T.: genetic analysis, interpreting data, writing—original draft preparation; G.B & K.T.A.: data curation, review and editing; M.B.; supervision, review and editing. All authors have read and agreed to the current version of the manuscript.

## Data Availability

All data produced in the present study are available upon reasonable request to the authors.

