## Supplementary figures and images for "Expanding the mutational spectrum of *GCK* in Turkish pediatric population"

### Supplementary Figure 1

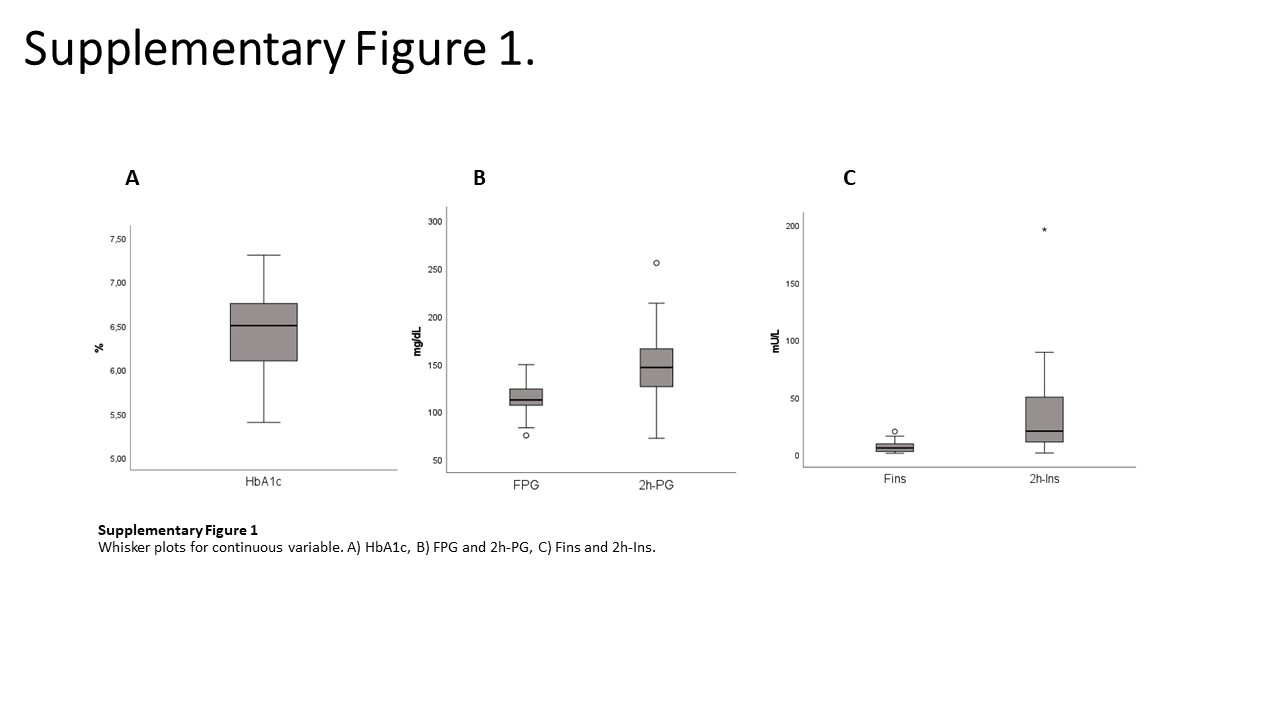

### Supplementary Figure 2

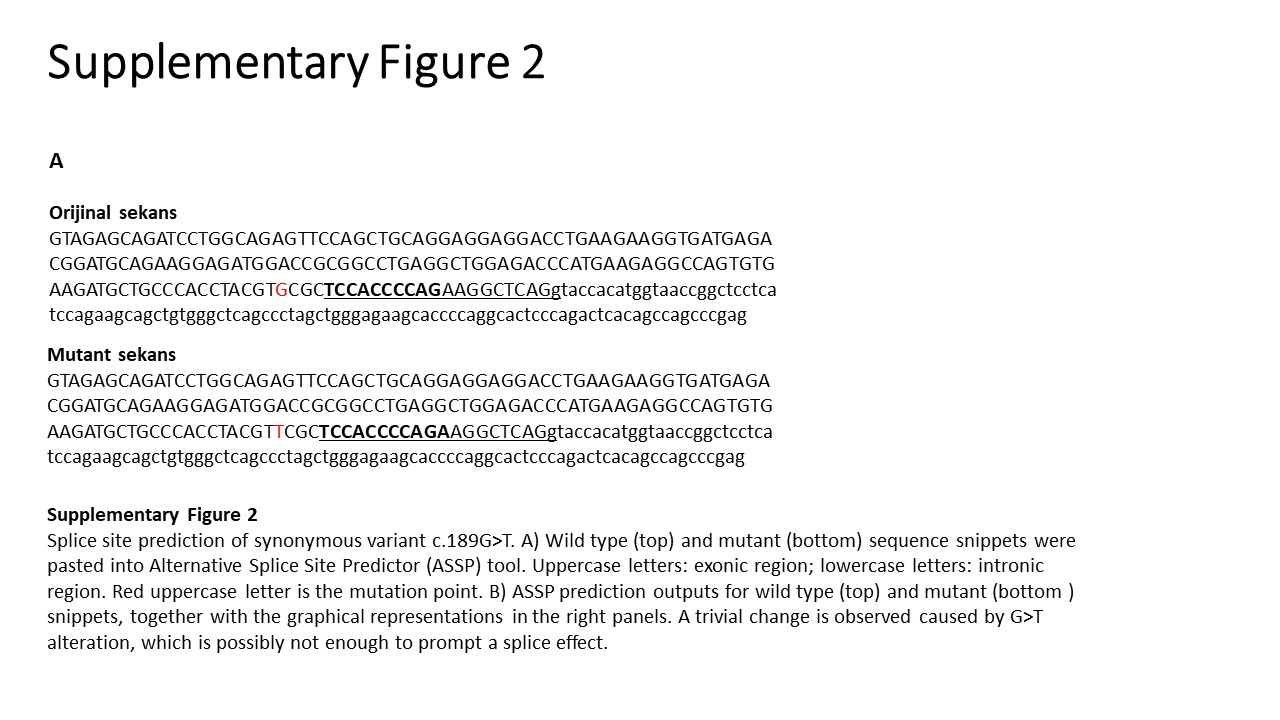

### Supplementary Figure 2

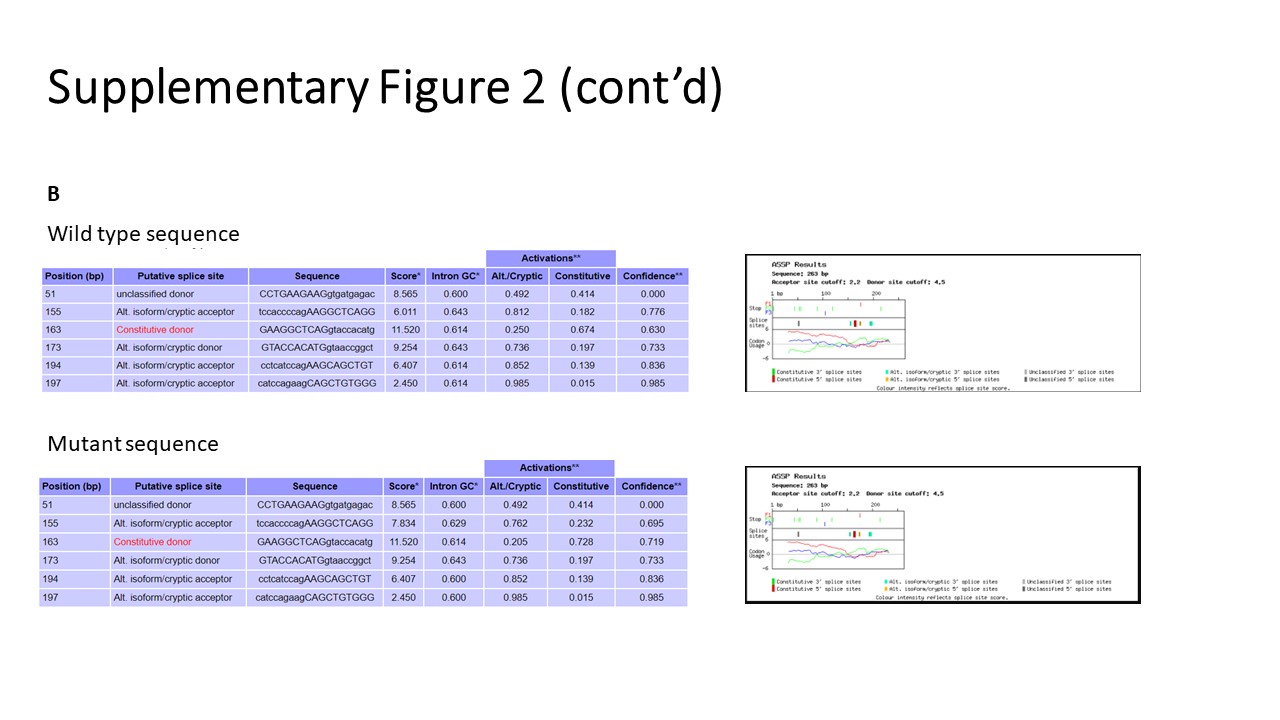
