## Supplementary Table 1 for "Expanding the mutational spectrum of *GCK* in Turkish pediatric population"

| **Supplementary Table 1 Literature search of variants detected in patients** | | | | | | | | | | |
| --- | --- | --- | --- | --- | --- | --- | --- | --- | --- | --- |
| **Reported/**  **Novelty** | **DNA** | **Protein** | **Times**  **detected**  **in this series** | **Previously reported**  **in Turkish**  **population** | **dbSNP** | **Clinvar (Accession)** | **HGMD** | **Functional**  **study** | **Reported defect(s)** | **Reporting researcher** |
| Reported | c.1342G>A | Gly448Ser | 1 | **no** | none | none | CM1916258 | yes | Protein instability and low protein yield | Li et al., 2022 |
| Reported | c.112C>T | Gln38* † | 2 | ‡yes | rs878853246 | P ** (VCV000236192.4) | none | no |  |  |
| Reported | c.950A>C | His317Pro | 1 | yes | none | none | none | no |  |  |
| Reported | c.1178T>C | Met393Thr | 9 | yes | none | none | CM096945 | yes | Altered enzyme stability and kinetics | Raimondo et al., 2014 |
| Reported | c.130G>A | Gly44Ser | 1 | yes | rs267601516 | P/LP (**) (VCV000076898.19) | CM013265, CM096811 | yes | Altered enzyme kinetics | Wang et al., 2019 |
| Reported | c.565A>G | Ile189Val | 1 | yes | rs757978639 | none | CM2113973 | no |  |  |
| Reported | c.208+3A>T | none | 6 | yes | none | none | CS1610081 | no |  |  |
| Reported | c.349G>C | Gly117Arg | 1 | **no** | none | none | none | no |  |  |
| Reported | c.863+5G>A | none | 1 | **no** | none | none | CS075156 | yes | Exon deletion | Bouvet et al., 2023 |
| Reported | c.214G>A | Gly72Arg | 1 | yes | rs193922289 | P* (VCV000976334.1) | CM023383 | yes | Altered enzyme stability and kinetics | Raimondo et al., 2014 |
| Reported | c.1292C>A | Thr431Lys | 1 | **no** | none | none | none | yes | Altered enzyme kinetics | Lin et., 2019 |
| **Novel** | c.830_831del | Val277fs*13 | 1 | **no** | none | none | none | no |  |  |
| Reported | c.106C>T | Arg36Trp | 1 | yes | rs762263694 | P/LP (**) (VCV000431973.21) | CM940823 | yes | Altered enzyme kinetics | Osbak et al., 2009 |
| Reported | c.572G>A | Arg191Gln | 1 | yes | rs886042610 | LP/P/VUS (*) (VCV000283358.20) | CM012120, CM096845 | †yes | Altered enzyme kinetics  (originally studied for R191W) | Wang et al., 2019 |
| Reported | c.186G>T | Val62Val | 1 | **no** | rs1244384220 | none | CD097020 | no |  |  |
| Reported | c.107G>C | Arg36Pro † | 1 | ‡yes | rs193922261 | LP(**) (VCV000036173.3) | CM096805 | no |  |  |
| Reported | c.454T>C | Phe152Leu | 1 | yes | none | none | CM064011, CD075428,  CM096830 | no |  |  |
| Reported | c.188G>A | Arg63His | 1 | **no** | rs746444094 (VUS) | none | none | no |  |  |
| Reported | c.793G>A | Glu265Lys | 1 | yes | rs104894011 | P** (VCV000447419.18) | CM984218, CM930303 | yes | Altered enzyme stability and kinetics | Osbak et al., 2009 |
| Reported | c.645C>A | Tyr215* | 1 | **no** | rs144723656 | P/LP** (VCV000453007.10) | CM004359, CM199048 | no |  |  |
| **Novel** | c.377T>A | Ile126Asn | 1 | **no** | none | none | none | no |  |  |
| Reported | c.667G>A | Gly223Ser | 1 | yes | rs1360415315 | P(**) (VCV000435306.18) | CM012123, CM096857 | yes | Altered enzyme kinetics | Valentinova et al., 2012 |
| Reported | c.46-1G>A | none † | 1 | ‡yes | none | none | CS1514535 | no |  |  |
| Reported | c.149A>C | His50Pro | 1 | **no** | none | none | CM012105, CM096812 | †yes | Altered enzyme kinetics  (originally studied for H50D) | Raimondo et al., 2014 |
| Reported | c.1079C>G | Ser360Trp † | 2 | ‡yes | none | none | CM970640 | no |  |  |
| Reported | c.401T>C | Leu134Pro | 1 | **no** | none | none | CM012112 | no |  |  |
| Reported | c.758T>G | Val253Gly † | 1 | ‡yes | rs193921400 | LP/LRA/VUS* (VCV000036253.4) | CM096877, CM1714218 | no |  |  |
| **Novel** | c.467_483+6del | none | 1 | **no** | none | none | none | no |  |  |
| ‡Reported only in Turkish patients. †These amino acids were not investigated directly but a different amino acid at this position was assayed. | | | | | | | | | | |
